# Brownfield land and emergency hospital admissions in England: a small-area ecological study using contamination hazard score as an exposure proxy

**DOI:** 10.64898/2026.10.30.26366956

**Authors:** Weiyi Wang, Garyfallos Konstantinoudis, Darren Beriro, Sarah Dack, Bethan Davies, Yunzhe Liu, Robie Kamanyire, Daniela Fecht

## Abstract

**Background:** Empirical evidence on the health impacts of brownfield land – previously developed land that may be contaminated – remains limited, mainly due to poorly defined exposures and pathways. Here we assessed associations between an area-level contamination hazard score and emergency hospital admissions across England, using the score as a proxy for the potential contamination associated with brownfield land.

**Methods:** We used Lower-layer Super Output Area (LSOA)-level population-weighted contamination hazard scores in Bayesian hierarchical models to examine the risk of emergency hospital admission (age- and sex-standardised, 2010–2019) with a primary diagnosis of i) all causes (excluding external causes), ii) respiratory diseases, and iii) cardiovascular diseases (CVD). Models were adjusted for deprivation, population density, tobacco expenditure, green space, urbanicity, and ethnicity, and included spatially structured and unstructured random effects to account for residual spatial autocorrelation between neighbouring areas. Stratified analyses assessed potential effect modification by deprivation, urban/rural classification, and region.

**Results:** We included 42,875,231 admissions for all natural causes, 6,657,050 for respiratory diseases, and 4,393,461 for CVD across 32,844 LSOAs in England. LSOAs with higher contamination hazard scores showed a non-linear decreasing trend in relative risk (RR) for all health outcomes. Spatial variation was evident, with stronger negative associations in urban and the least deprived LSOAs, while rural and more deprived LSOAs showed weaker associations or greater uncertainty. Region-specific analyses demonstrated inverse associations for all outcomes in London and for all natural causes and respiratory admissions in the North West and East of England. Other regions showed no consistent pattern across outcomes.

**Conclusion:** Higher contamination hazard scores were associated with lower emergency hospital admission risks. This suggests that areas with a greater legacy of potentially contaminating land uses do not necessarily experience poorer contemporary population health. The inverse associations may partly reflect subsequent remediation and urban regeneration, although the underlying mechanisms require further investigation.

**Highlights:**

- A national brownfield study used contamination hazard scores as an exposure proxy.
- Higher contamination hazard scores were linked to lower emergency admission risks.
- Inverse associations were strongest in urban areas, least deprived areas and London.
- Lower risks may reflect redevelopment, not a protective effect of brownfield land.

## 1. Introduction

Brownfield land, or previously developed land, has become an important component of urban regeneration plans in the United Kingdom (UK). The UK is one of the most urbanised countries in the world. Currently, over 80% of the population live in urban areas, with a projected steady and continuous increase over the next few decades (United Nations, 2025). This predominantly urban growth is expected to increase demand for housing, infrastructure and services. It has also created pressure to limit urban sprawl and protect greenfield land from development. In 2024, the UK government set the target to build 1.5 million new houses in England by 2029 (HM Government, 2024). Policies to support this target include a ‘default yes’ approach to brownfield development. Funding streams such as the Brownfield Land Release Fund provide support for local authorities to unlock underused or derelict brownfield sites for housing-led redevelopment (Local Government Association, 2026).

Redeveloping brownfield land is an important part of sustainable urban planning. While greenfield (i.e. undeveloped green land, generally located on the suburban fringes) development can contribute to habitat and ecosystem degradation (Eigenbrod et al., 2011; Seto et al., 2012), brownfield sites are often located within urban areas, and many are already connected to transport, utilities and other infrastructure (Jacek et al., 2022). Their redevelopment may also support local economic renewal by supporting growth of the local labour market, improve local environments and reconnect fragmented communities (He et al., 2024; Hou et al., 2023). Brownfield redevelopment may therefore offer economic and environmental advantages over greenfield development.

Depending on their previous use, brownfield sites may, however, be heavily contaminated. Former industrial or commercial activities may leave hazardous substances (e.g. heavy metals, hydrocarbons, solvents, and asbestos) in soil, groundwater, and/or buildings (Hellawell and Hughes, 2021; Jacek et al., 2022). Site conditions may also change over time through dereliction, vandalism, fires, fly-tipping, or demolition and remediation works, which can alter the contaminant profiles and potentially mobilise pollutants (Wang et al., 2023). Local residents may be exposed to these contaminants through multiple pathways, including inhalation of vapours or contaminated dust, ingestion of contaminated soil, dust or water, and direct contact with contaminated materials (Environment Agency, 2025). Vacant or derelict land can contribute to neighbourhood blight, community stigma and poor perceived environmental quality. These factors may increase psychological stress and reduce wellbeing (Bambra et al., 2015; Garvin et al., 2013). Such sites may also attract vandalism and illegal waste dumping, with accumulated combustible waste posing a fire risk (London Borough of Havering, 2023). Therefore, brownfield land has a dual relevance for public health research. It may represent a potential source of contamination associated with historical industrial or commercial activities which can adversely affect human health. It may also act as a marker of broader social, environmental and urban change.

Despite growing policy interest, epidemiological evidence on brownfield land and health remains limited. A recent systematic review identified only six relevant studies. These studies reported associations with poorer self-reported health, higher mortality, birth defects, elevated serum metal levels and immune ageing (Wang et al., 2023). Two more recent studies have since extended the evidence base, reporting positive associations between lead-contaminated brownfield land and birth defects, and area-level brownfield frequency and poor self-reported mental health (Kulali et al., 2025; Slawsky et al., 2024).

Existing studies vary in their exposure assessment, study design and outcome measures. There is also limited understanding of whether associations vary by socioeconomic deprivation and geographical context, despite brownfield land being socially and spatially patterned. Investigating the health implications of brownfield land is challenging because the diverse exposures and pathways are difficult to capture, especially at the population level. For this study, we used a spatially resolved contamination hazard score as an area-level proxy for potential contamination. We examined associations between the score and emergency hospital admissions in England using an ecological study design, with stratified analyses to assess effect modification by deprivation and geographical settings. The study therefore addresses potential contamination as one health-relevant aspect of brownfield land, rather than the health effects of brownfield extent or actual contaminant exposure events.

## 2. Materials and methods

### 2.1 Health outcomes

We obtained emergency hospital admission counts for each Lower-layer Super Output Area (LSOA) in England from NHS Digital Hospital Episodes Statistics (HES) held at the UK Small Area Health Statistics Unit (SAHSU) for 2010 to 2019. LSOAs are census dissemination areas, typically containing 1,000–3,000 usual residents, which we used to represent neighbourhoods in this study (ONS, 2022). We examined primary diagnoses for admissions due to all causes (excluding external causes; International Statistical Classification of Diseases and Related Health Problems, 10^th^ revision [ICD-10]: A00–R99), respiratory diseases (ICD-10: J00–J99), and cardiovascular diseases (ICD-10: I00–I99). These outcomes were selected as broad indicators of population health.

Hospital admission counts were age- and sex-standardised using indirect standardisation. Population data were taken from the Office for National Statistics (ONS) mid-year population estimates (ONS, 2020). Briefly, for each age-sex stratum, a reference rate was calculated as the total number of admissions divided by the total population in that stratum across the reference population (all of England). These reference rates were applied to the age-sex population of each LSOA to obtain the expected number of admissions per stratum, under the assumption that each LSOA experienced the same stratum-specific rates as the reference population. Observed (*O*) and expected (*E*) counts were then summed across all age-sex strata to give the total observed and expected number of admissions per LSOA:

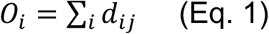

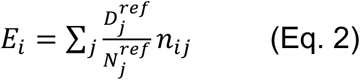

where *i* indexes LSOAs and ( indexes age-sex strata; *d_i_*_j_ and *n_i_*_j_ are the observed admissions and population in stratum ( of LSOA *i* ; and 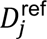 and 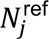 are the total admissions and population in stratum (of the reference population, respectively.

The study was covered by national research ethics approval from the London–South East Research Ethics Committee (Reference 22/LO/0256). Data access was covered by the Health Research Authority Confidentiality Advisory Group under section 251 of the National Health Service Act 2006 and the Health Service (Control of Patient Information) Regulations 2002 (Reference 20/CAG/0028).

### 2.2 Exposure

Contamination is one of the key health-relevant concerns associated with brownfield land. We used a contamination hazard score as an area-level proxy for potential land contamination associated with brownfield sites. Contamination hazard scores for England were obtained from Groundsure, an independent environmental consulting company specialising in environmental and climate data. Scores were available only for land with previous industrial use recorded on historical Ordnance Survey maps. Such land may remain in industrial use, have become vacant, or now accommodate housing, offices, car parks or open space. Its coverage across England and spatially detailed contamination hazard information provided a basis for examining area-level health associations relevant to potential contamination associated with previous land uses.

The scores were provided on a hexagonal grid with a cell area of 0.25 ha. The scoring methodology uses Geographic Information System-based Multi-Criteria Decision Analysis (GIS-MCDA) to integrate environmental and geological spatial data and generate scores for contamination and geotechnical hazards within a source-pathway-receptor (S-P-R) framework originally developed by the British Geological Survey (Hammond et al., 2024). Within the framework, potential contaminant source datasets included Historical Land Use (Groundsure and Ordnance Survey), Recorded Pollution Incidents and Historical Landfills (Environment Agency); pathway datasets included Boreholes (British Geological Survey) and Mine Entries (Mining Remediation Authority); and receptor datasets included Groundwater Vulnerability (Environment Agency/British Geological Survey) and Surface Water Features (Ordnance Survey). A weighted sum model (WSM) was used to generate a score for each S-P-R component. These component scores were then combined as follows:

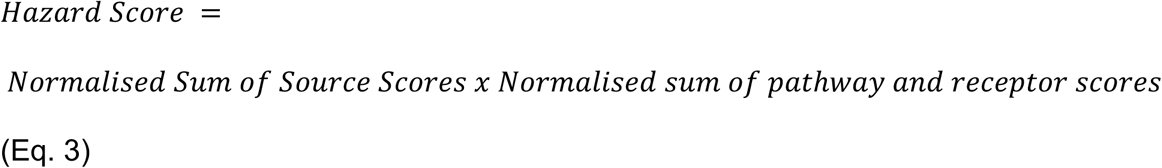

The contamination hazard scores were overlaid with the 2011 LSOA boundaries and a point layer containing population count data for each postcode centroid (average of 12–15 households), both from the ONS Census 2011. The resulting point layer included the contamination hazard score, population count, and the corresponding LSOA for each postcode centroid. For each LSOA, we calculated a population-weighted mean contamination hazard score by multiplying the score at each postcode centroid by the corresponding postcode population, summing these products across all postcodes within the LSOA, and dividing by the total population of those postcodes:

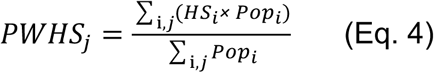

where *PwHS*_j_ is the population-weighted contamination hazard score for LSOA *j*, *HS_i_* is the contamination hazard score at the centroid of postcode i, and *Pop_i_* is the population count for postcode i. The summations include all postcodes i within LSOA *j*.

The resulting LSOA-level population-weighted contamination hazard score (hereafter referred to as the contamination hazard score) was linked to the observed and expected hospital admission counts for each LSOA. Associations were examined using a cross-sectional ecological design, with LSOAs as the units of analysis.

The spatial analysis was conducted in R statistical software (version 4.4.2) using the sf package.

### 2.3 Confounders

We identified potential confounders a priori based on the literature, including area-level socioeconomic deprivation, population density, smoking, greenness, urbanicity, non-White ethnicity and traffic-related air pollution.

We used the income domain of the 2019 Index of Multiple Deprivation (IMD) as a relative measure of LSOA-level socioeconomic deprivation. The IMD is a composite area-level measure of deprivation consisting of several domains (McLennan et al., 2019). The income domain is the proportion of the population claiming income-related benefits due to being out of work or having low earnings. Income deprivation was categorised into quintiles, representing the least deprived 20% of LSOAs (quintile 1) to the most deprived 20% of areas (quintile 5). We used the 2019 mid-year population estimates from the Office for National Statistics (ONS) to calculate population density as the number of residents per unit area for each LSOA. Individual-level smoking information for England is not available. We therefore used LSOA-level tobacco expenditure (£ per week per person aged ≥16 years) sourced from CACI Ltd. (©2024) as an area-level proxy for smoking. To account for greenness, we included the normalised difference vegetation index (NDVI), derived from Sentinel-2 images via Google Earth Engine for the year 2019 for each LSOA. Images with less than 10% cloud cover were retained, and mean NDVI was calculated across images. The resulting NDVI layer was exported at 100 m resolution and averaged within each LSOA. Population density, tobacco expenditure, and NDVI were used as continuous variables. We also included a binary covariate on urbanicity (urban/rural) based on the rural urban classification provided by ONS (ONS, 2016). Additionally, we obtained ethnicity data from the 2011 Census and calculated the percentage of residents recorded as non-White in each LSOA. Annual average concentrations of nitrogen dioxide (NO₂), a traffic-related air pollution indicator modelled using hybrid land use regression modelling at 25 m resolution (Wang et al., 2022), were population-weighted to the LSOA level.

### 2.4 Statistical analysis

We used a Bayesian hierarchical modelling approach to assess the associations between population-weighted contamination hazard scores and age- and sex-standardised emergency hospital admissions in England. Spatial dependence was modelled using the BYM2 parameterisation of the Besag-York-Mollié (BYM) model (Besag et al., 1991; Riebler et al., 2016). Models also included a region-level random intercept to account for additional variation between regions. The contamination hazard score was modelled using a second-order random walk to allow for a non-linear association with hospital admissions. The outcome was the observed number of hospital admissions (*Y_k_*) in each LSOA (*k*), assumed to follow a Poisson distribution. The expected number of admissions (*e_k_*) was calculated using age-sex-specific admission rates. Its natural logarithm, *log*(*e_k_*), was included as an offset to account for differences in population size and demographic composition between LSOAs. Population density and contamination hazard scores were log-transformed, with a small constant added to the scores to accommodate zero values. We developed fully adjusted models as follows:

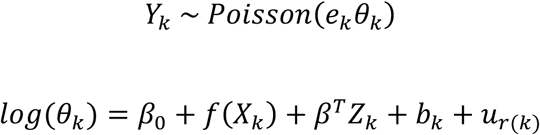

Here, J*_k_* represents the underlying relative risk of hospitalisation in LSOA *k*. *β*_0_ is an intercept; *X_k_* denotes the grouped, log-transformed population-weighted contamination hazard score, modelled using a flexible non-linear function, *f*(*X_k_*), specified as a second-order random walk. *β* is the vector of regression coefficients, and *Z_k_* is the vector of covariate values for LSOA *k*. The covariates include (1) population density, (2) income deprivation quintiles, (3) tobacco expenditure, (4) NDVI, (5) rural/urban classification, and (6) ethnicity. NO_2_ was not included in the main models because it was strongly correlated with population density and ethnicity (Pearson’s *r* = 0.70). The term *b_k_* represents a BYM2 spatial random effect, which combines spatially structured and unstructured components to account for residual spatial variation between LSOAs. The term *u_r_*_(*k*)_ represents an independent region-level random intercept, where *r*(*k*) identifies the region containing LSOA *k*.

Exposure-response curves were centred post-estimation at a common reference, defined as the median contamination hazard score across all LSOAs, so that relative risks were interpreted relative to this value and were comparable across outcomes and stratified analyses. We explored possible effect modification in stratified analyses by income deprivation quintile, urban/rural classification, and region (nine regions of England).

As a sensitivity analysis, we fitted additional models in which the contamination hazard score was entered as a categorical exposure rather than as a continuous term to assess whether the findings were sensitive to the modelling approach for the exposure. We used two categorisations: i) three categories, with zero as a separate category and the 90^th^ percentile as a cut-off; and ii) four categories, with zero as a separate category and the median and 95^th^ percentile as cut-offs. In addition, we repeated the analyses using linear regression models to assess the robustness of the findings to the choice of modelling framework.

All models were fitted using the R package INLA (Rue et al., 2009).

## 3. Results

All 32,844 LSOAs in England were included in the analysis. Figure 1 shows the spatial variation in the population-weighted contamination hazard score at the LSOA level across the nine regions of England. A total of 1,998 LSOAs (6.1%) had a contamination hazard score of zero. The LSOA-level contamination hazard scores ranged from 0 to 0.69, with most LSOAs falling within the >0–0.05 category (79%). Distinct hotspots were observed in several urban and industrial settings. The highest scores were concentrated in London, where contiguous clusters of LSOAs in east London exceeded 0.2, with some exceeding 0.4 in places. Additional hotspots were observed in parts of the West Midlands, the North West, and Yorkshire and the Humber. These hotspots broadly coincided with urban areas with a history of industrial activity, consistent with the legacy of industrial land use (2002).

**Figure 1.**
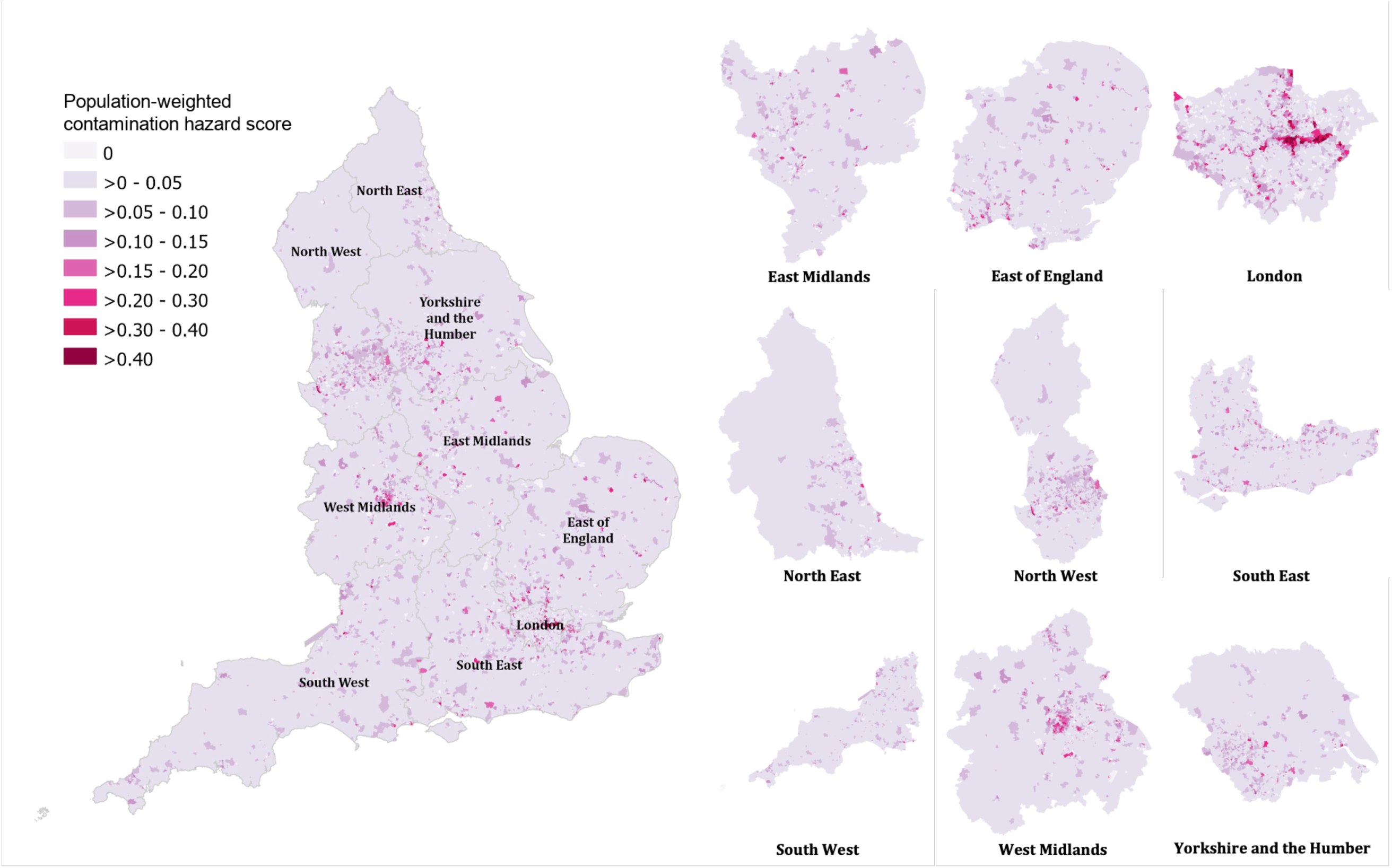
Spatial distribution of population-weighted contamination hazard scores at the LSOA level across the nine regions of England.

Spatial variation in the six covariates at the LSOA level across England and boundaries of the nine regions of England are shown in Supplementary Fig. S1. Urban areas were generally characterised by higher population density and income deprivation, a higher proportion of non-White residents and lower NDVI, while tobacco expenditure showed a north-south gradient rather than a clear urban-rural pattern. Correlations between the contamination hazard score and potential confounders were generally weak (Pearson’s *r* = –0.15 to 0.14, Supplementary Fig. S3).

Table 1 summarises the distribution of emergency hospital admissions by income deprivation quintile, urban/rural classification, and region for all natural causes, respiratory disease, and CVD. In total, there were 42,875,231 emergency hospital admissions due to all natural causes, including 6,657,050 due to respiratory disease and 4,393,461 due to CVD. A clear graded pattern in admission counts and shares was observed across deprivation quintiles, with a greater proportion of admissions occurring in more deprived areas. Hospital admissions were predominantly concentrated in urban areas (range across outcomes: 81.3%–85.1%), reflective of the urban population in England (83.2%). For CVD, however, rural residents accounted for a slightly higher proportion of admissions (18.7%) than of the population (16.8%). The North West accounted for the largest share of hospital admissions across all three outcomes. Regional patterns broadly reflected regional population distributions, although the North West had a slightly higher proportion of hospital admissions relative to its population size, whereas London showed the opposite. The distributions of continuous area-level covariates across LSOAs are summarised in Supplementary Table S1.

**Table 1.**
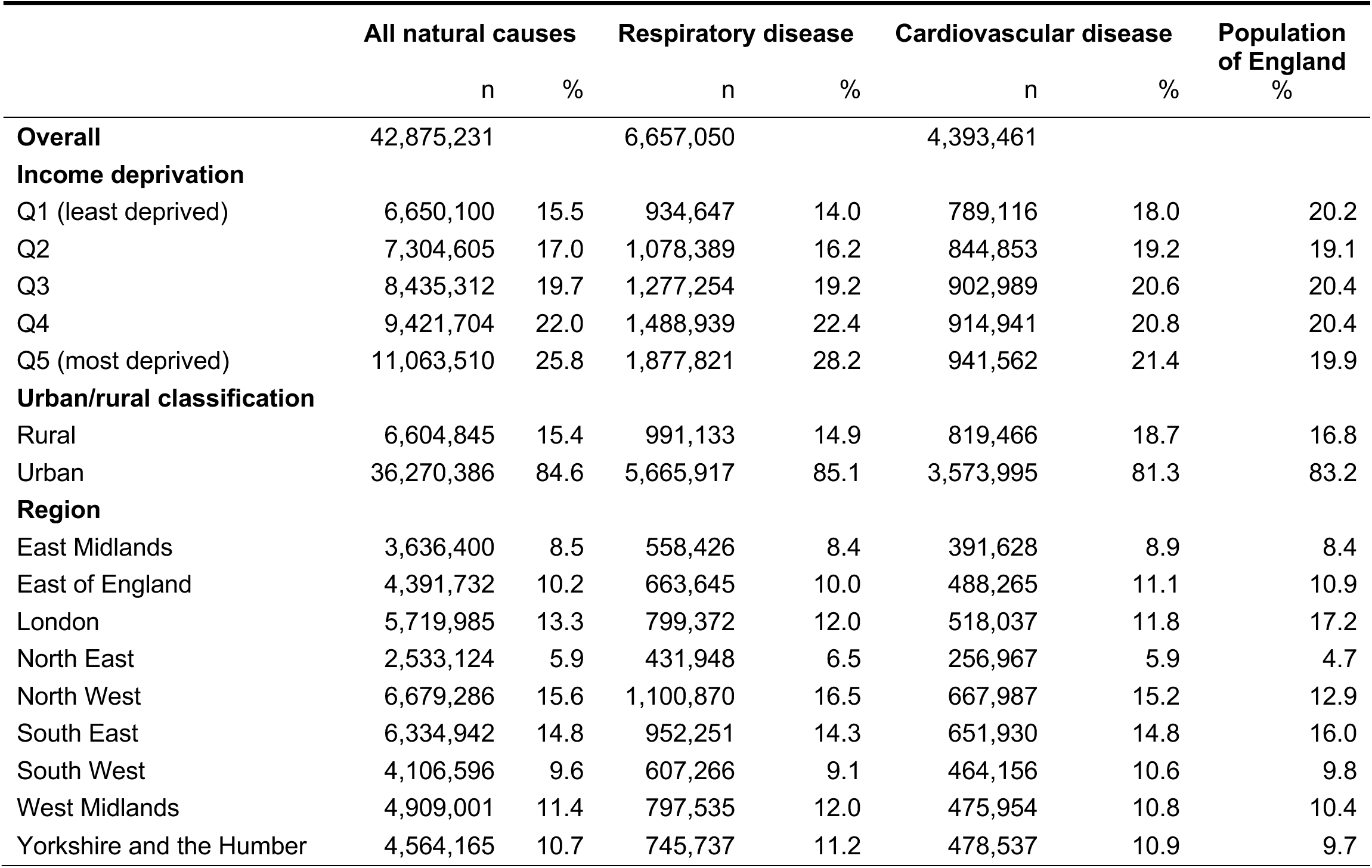
Distribution of emergency hospital admissions and the population of England by socioeconomic and geographic characteristics.

|  | **All natural causes** |  | **Respiratory disease** |  | **Cardiovascular disease** |  | **Population of England** |
|---|---|---|---|---|---|---|---|
|  | n | % | n | % | n | % | % |
| **Overall** | 42,875,231 |  | 6,657,050 |  | 4,393,461 |  |  |
| **Income deprivation** |  |  |  |  |  |  |  |
| Q1 (least deprived) | 6,650,100 | 15.5 | 934,647 | 14.0 | 789,116 | 18.0 | 20.2 |
| Q2 | 7,304,605 | 17.0 | 1,078,389 | 16.2 | 844,853 | 19.2 | 19.1 |
| Q3 | 8,435,312 | 19.7 | 1,277,254 | 19.2 | 902,989 | 20.6 | 20.4 |
| Q4 | 9,421,704 | 22.0 | 1,488,939 | 22.4 | 914,941 | 20.8 | 20.4 |
| Q5 (most deprived) | 11,063,510 | 25.8 | 1,877,821 | 28.2 | 941,562 | 21.4 | 19.9 |
| **Urban/rural classification** |  |  |  |  |  |  |  |
| Rural | 6,604,845 | 15.4 | 991,133 | 14.9 | 819,466 | 18.7 | 16.8 |
| Urban | 36,270,386 | 84.6 | 5,665,917 | 85.1 | 3,573,995 | 81.3 | 83.2 |
| **Region** |  |  |  |  |  |  |  |
| East Midlands | 3,636,400 | 8.5 | 558,426 | 8.4 | 391,628 | 8.9 | 8.4 |
| East of England | 4,391,732 | 10.2 | 663,645 | 10.0 | 488,265 | 11.1 | 10.9 |
| London | 5,719,985 | 13.3 | 799,372 | 12.0 | 518,037 | 11.8 | 17.2 |
| North East | 2,533,124 | 5.9 | 431,948 | 6.5 | 256,967 | 5.9 | 4.7 |
| North West | 6,679,286 | 15.6 | 1,100,870 | 16.5 | 667,987 | 15.2 | 12.9 |
| South East | 6,334,942 | 14.8 | 952,251 | 14.3 | 651,930 | 14.8 | 16.0 |
| South West | 4,106,596 | 9.6 | 607,266 | 9.1 | 464,156 | 10.6 | 9.8 |
| West Midlands | 4,909,001 | 11.4 | 797,535 | 12.0 | 475,954 | 10.8 | 10.4 |
| Yorkshire and the Humber | 4,564,165 | 10.7 | 745,737 | 11.2 | 478,537 | 10.9 | 9.7 |

Relative risks (RRs) were estimated against a reference contamination hazard score of 0.019, corresponding to the median score across all LSOAs. Across the three outcomes, RRs remained close to the null at very low scores, with no clear evidence of increased risk. (Fig 2a). Above the reference value, associations showed a broadly monotonic decline with increasing scores. For hospital admissions due to all natural causes, RR fell below the null and reached statistical significance from a contamination hazard score of 0.13, with reductions of 1.7% at a contamination hazard score of 0.15, reaching 15.8% at the highest observed score. Hospital admissions due to respiratory diseases followed a similar pattern, with reductions of up to 12.1% at the highest contamination hazard score. Associations for hospital admission due to CVD declined more gradually, remaining close to the null up to a score of 0.20, before decreasing more steeply, with statistically significant reductions of up to 12.5%. Credible intervals overlapped substantially across outcomes and widened at higher scores, reflecting data sparsity at the upper end of the exposure distribution (i.e. a small number of LSOAs had high scores). Unadjusted models showed a similar pattern to the fully adjusted models, with higher RRs at low scores and stronger inverse associations at higher scores (Supplementary Fig. S2).

**Figure 2.**
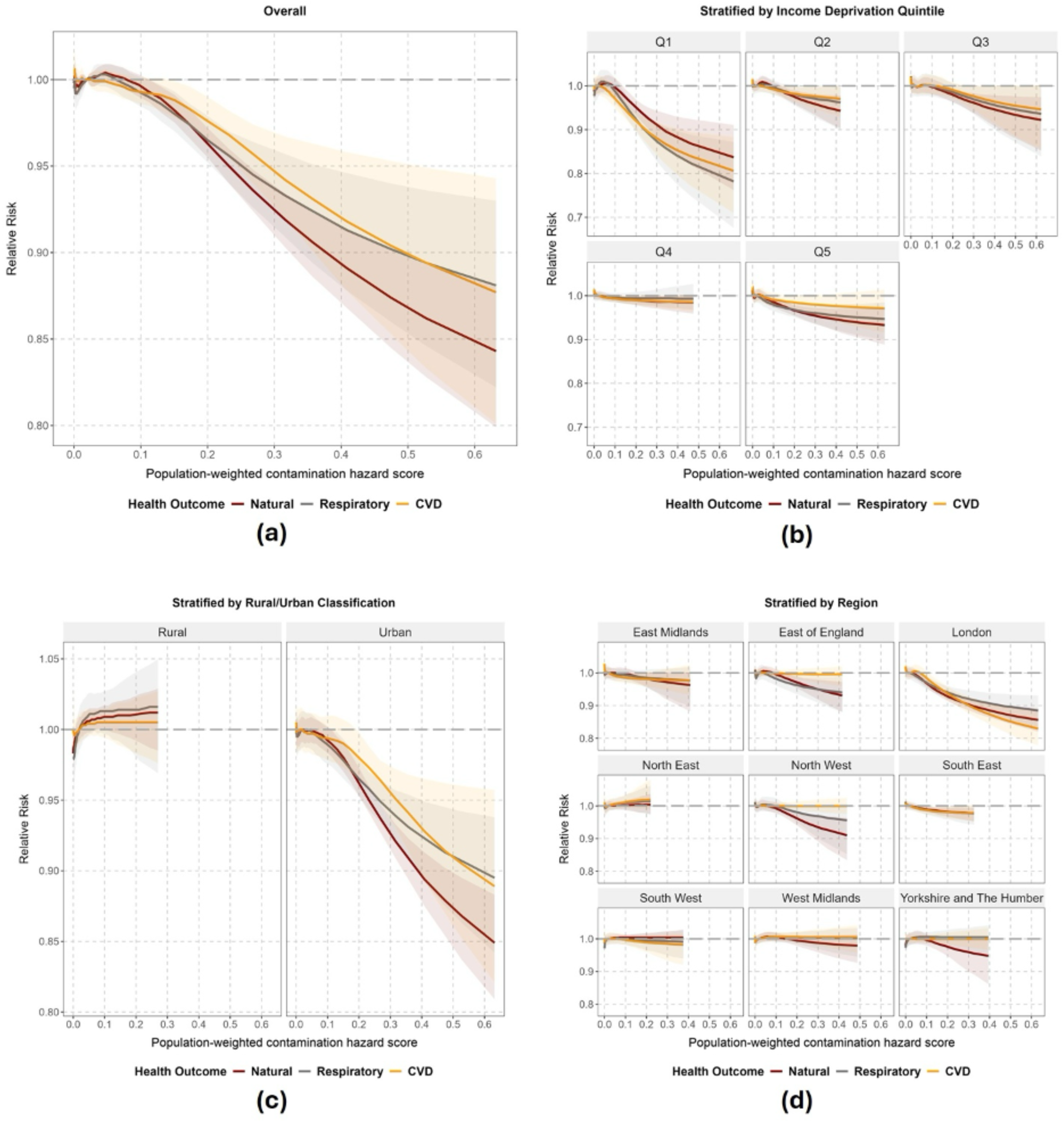
Posterior median relative risks and 95% credible intervals estimated from a Bayesian hierarchical model, showing associations between LSOA-level population-weighted contamination hazard scores and hospital admissions for all natural causes (Natural), respiratory diseases (Respiratory), and cardiovascular diseases (CVD): a) overall association, and stratified analyses by b) income deprivation quintile (Q1 – least deprived), c) urban/rural classification, and d) region. The dashed line at 1.0 represents the null (no effect).

When stratified by income deprivation quintile, the inverse association between contamination hazard score and hospital admissions varied across deprivation groups (Fig. 2b). The strongest associations were observed in the least deprived areas (Q1), particularly for respiratory admissions, and attenuated across more deprived quintiles. In Q1, risks were largely centred around the null at very low scores, with a small yet non-significant initial increase across all three health outcomes. RRs then declined, reaching statistical significance from around a score of 0.15 (approximately the 97^th^ percentile of the LSOA-level score distribution) for all natural causes and respiratory causes, and from around 0.09 for CVD. Across the statistically significant range, estimated risk reductions varied substantially, from 2.4% to 16.3% for hospital admissions due to all natural causes, 3.0% to 22.0% for respiratory disease-related hospital admissions, and 2.6% to 19.7% for CVD-related hospital admissions. By contrast, associations in Q2–Q4 were weak and largely centred around the null, with slight downward trends and wider credible intervals at higher contamination hazard scores. In Q5, modest inverse associations were observed for all natural causes and respiratory disease-related hospital admissions at higher contamination hazard scores, although estimates at the upper end were highly uncertain, with only two LSOAs at the upper end of the score distribution.

Stratification by urban/rural classification showed a clear contrast between LSOAs classified as urban versus rural (Fig. 2c). In rural areas, associations for all three outcomes remained close to the null and not statistically significant across the exposure range. Rural LSOAs also had generally lower contamination hazard scores and a narrower exposure range than urban LSOAs, limiting comparisons at higher scores. By contrast, the exposure-response patterns in urban areas broadly mirrored the overall associations, with slightly increased risks at low contamination hazard scores followed by a progressive decline at higher scores. This similarity suggests that the overall pattern may have been largely driven by urban areas.

Stratification by region showed substantial regional heterogeneity in associations between contamination hazard scores and hospital admissions (Fig. 2d). The strongest inverse associations were observed in London. RRs generally declined with increasing contamination hazard scores above the reference value, with statistically significant reductions observed from around a contamination hazard score of 0.10 across all hospital admission outcomes. Estimated reductions ranged from 3.7% to 14.5% for hospital admissions due to all natural causes, 2.7% to 11.6% for respiratory disease-related hospital admissions, and 2.1% to 17.1% for CVD-related hospital admissions. A comparable but weaker inverse gradient was observed in the North West and East of England for hospital admissions due to all natural causes and respiratory diseases, while associations for CVD-related hospital admissions in both regions were largely flat and remained close to the null. In the other regions, associations were generally flat and/or centred around the null, with no consistent exposure-response pattern across health outcomes.

In sensitivity analyses, we examined whether the exposure-response pattern observed in the continuous models was influenced by the skewed distribution of the contamination hazard score towards low scores (Supplementary Figure S4). We first divided the score into four categories: zero (0; reference), low (>0 to 0.019), moderate (>0.019 to 0.121) and high (>0.121), where 0.019 and 0.121 corresponded to the median and 95^th^ percentile, respectively. RRs were close to 1 for all outcomes in the low and moderate categories (Supplementary Table S2). Compared with the reference group, the high category was associated with lower RRs of hospital admissions due to all natural causes (RR: 0.975, 95% CrI: 0.966–0.984), respiratory causes (RR: 0.982, 95% CrI: 0.968–0.997), and CVD causes (RR: 0.980, 95% CrI: 0.968–0.992).

We then fitted a three-category model comprising zero (0; reference), non-zero scores up to the 90^th^ percentile (>0 to 0.083), and scores above the 90^th^ percentile (>0.083), thereby including more LSOAs in the highest-score group. Results were very similar: RRs remained close to 1 for non-zero scores up to the 90^th^ percentile, while scores above the 90^th^ percentile were associated with lower RRs across all outcomes (Supplementary Table S2). Overall, results of the categorical analyses were consistent with those of the continuous models, showing lower RRs for areas with higher contamination hazard scores. The inverse association at higher scores was also observed in a linear regression sensitivity analysis without spatial random effects. Compared with LSOAs with zero contamination hazard scores, coefficients for the moderate- and high-score categories were negative across all three outcomes, with the largest differences in the high-score category (Supplementary Table S3).

## 4. Discussion

In this national small-area analysis, areas with higher contamination hazard scores were generally associated with lower risks of emergency hospital admissions for all natural causes, respiratory diseases, and CVD, while there was no clear evidence of increased risk at very low scores. The strength of the association varied across population subgroups and geographical contexts. The sensitivity analyses also showed lower risks in categories above the 90^th^ or 95^th^ percentile, suggesting that the inverse association was unlikely to be an artefact of modelling the contamination hazard score as a continuous exposure. The inverse association was most pronounced in the least income-deprived areas (Q1), in urban settings, and in London.

The observed inverse associations require cautious interpretation and are unlikely to indicate a direct protective effect of potential contamination. Several contextual factors may contribute to this counterintuitive finding.

First, the contamination hazard score does not establish whether exposure pathways are active. Buildings, hardstanding and restricted access may limit people’s contact with contaminated soil and dust, even where contamination remains. Sites within areas with higher scores may have undergone remediation and development to provide green spaces, parks, community amenities or better-quality housing, potentially reducing contamination-related exposures and providing environmental and social benefits that may contribute to improved health outcomes (Hou et al., 2023). Although green space was included in the models, other changes associated with industrial decline and regeneration may not have been fully captured. Therefore, it is possible that contemporary health outcomes partly reflect the regenerated landscape rather than current exposure to contaminants.

Second, population mobility and selective migration may further explain the observed pattern. City centre and regenerated areas often attract younger, healthier, or more affluent residents who are more physically active, while more vulnerable population may move away or be displaced through gentrification and rising housing costs (Keddie and Tonkiss, 2010; Leccis, 2019). Such selective migration may alter the health profile of residents, potentially contributing to lower hospital admission risks (Norman et al., 2005). Adjustment for area-level income deprivation may not fully capture these changes in population composition.

Third, residual confounding may remain because differences in unmeasured factors, such as healthcare access, housing quality and local environmental amenities, may contribute to the observed association. Emergency admissions also reflect disease management, care-seeking behaviour and hospital admission practices, as well as underlying morbidity. Variation in these processes across urban and socioeconomic contexts may partly explain the observed associations.

These findings contrast with the adverse health associations reported in previous studies of brownfield land (Kulali et al., 2025; Slawsky et al., 2024; Wang et al., 2023). However, exposure definitions varied across this literature, including site counts, residential proximity and hazard-based scores incorporating historical land use. For example, Litt et al. (2002) classified census tracts using a hazard score incorporating historical land use and reported higher cancer mortality, including lung cancer mortality, in high-hazard compared with low-hazard areas. Differences in both exposure assessment and health outcomes limit direct comparability with the present study. The negative associations observed in the present study may therefore reflect differences in the exposure metric and in the health outcomes, geographical scales and study periods examined. Notably, the England-wide area-level study by Bambra et al. (2015) reported regional variation in associations between brownfield land and health. Their adjusted comparisons showed higher premature mortality in the North West and lower premature mortality in London than in the South East within corresponding brownfield categories. These findings concern differences between regions, whereas our London analysis examines associations with contamination hazard scores within the region.

This study, to our knowledge, is the first national small-area analysis in England to examine associations between contamination hazard scores and emergency hospital admissions. The study used a large, national dataset of emergency hospital admissions over multiple years, allowing for detailed stratified analyses. The spatial modelling framework accounted for underlying geographical structure, while regional analyses allowed exploration of heterogeneity across different urban and regional contexts. Additionally, unlike studies relying solely on brownfield proximity or density, the contamination hazard score provided an area-level indication of potential contamination based on source-pathway-receptor principles.

Several limitations should be considered. First, the contamination hazard score is an aggregate area-level indicator of potential contamination, rather than a direct measure of brownfield extent or individual measured contaminant concentrations and exposure. This means the scores do not establish the extent to which residents come into contact with contaminated soil, water, or air, potentially leading to exposure misclassification. Furthermore, aggregation at the LSOA level may mask local variation in contamination hazard scores and obscure associations affecting small populations living close to individual sites. Second, the ecological design limits the interpretation of area-level associations in terms of individual exposure and health risks. Third, residual confounding is likely, particularly from unmeasured area-level factors such as healthcare access, housing conditions, population mobility, and other environmental exposures. Fourth, the inverse associations observed may reflect structural or compositional differences between areas, and should not be interpreted as evidence that potential contamination is protective. These limitations are particularly relevant in highly urban and regenerated settings, where higher contamination hazard scores may partly reflect redevelopment processes rather than current contaminant exposure.

## 5. Conclusions

Our study found that higher population-weighted contamination hazard scores were associated with lower risks of emergency hospital admissions in England, with substantial variation by deprivation, urbanicity, and region. The associations were context-dependent and should not be interpreted as evidence that brownfield land is protective. The contamination hazard score may also reflect broader social and environmental conditions associated with historical land use and subsequent redevelopment. For brownfield redevelopment policy, historical contamination potential should therefore be considered alongside current site conditions, remediation history, and changes in site and surrounding land use, when assessing potential health implications. Individual-level data and longitudinal designs pertaining to the role contaminated brownfield land plays in the health of local communities would help clarify the mechanisms underlying the observed area-level patterns and inform public health considerations in brownfield redevelopment, housing and urban planning policies.

## 7. Data availability

Data may be obtained from a third party and are not publicly available.

## 8. Acknowledgements

We extend thanks to Groundsure for their generous contribution of their commercial dataset, Groundscreen, for the purposes of our research. We also thank Hima Daby and Gajanan Natu for their help with data acquisition, storage, preparation and governance. The views expressed are those of the authors. This paper does not necessarily reflect the views of the National Institute for Health and Care Research or the Department of Health and Social Care. Hospital Episode Statistics (hospital admissions) data are copyright © 2026, re-used with the permission of NHS Digital. All rights reserved.

Includes data derived from source datasets owned by the British Geological Survey © UKRI, the Mining Remediation Authority, and Ordnance Survey. All rights reserved.

