## Supplementary Material for "Brownfield land and emergency hospital admissions in England: a small-area ecological study using contamination hazard score as an exposure proxy"

Figure S**1**. Spatial distribution of covariates at the LSOA level.

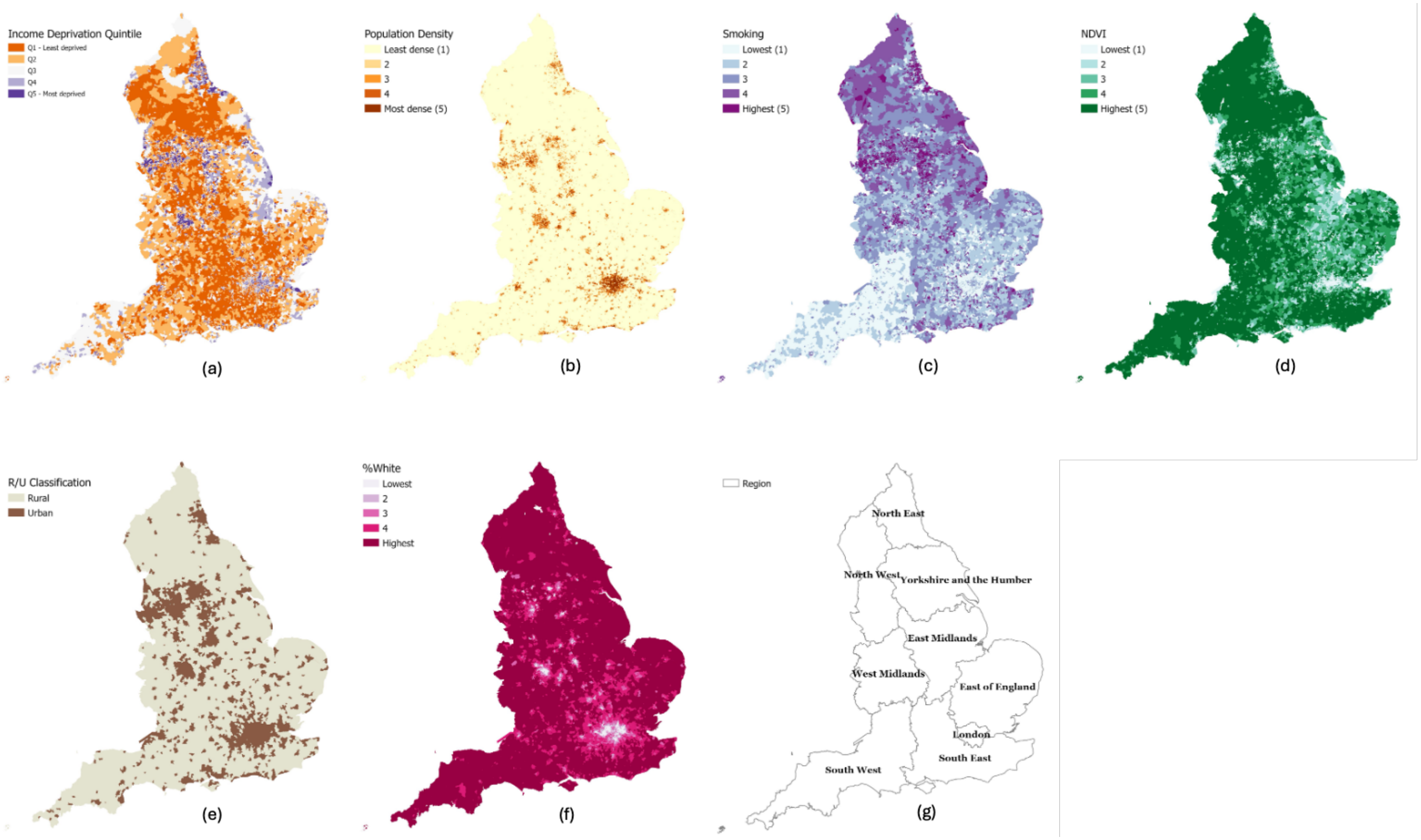

Note: (a) income deprivation quintile, with Q1 representing the least deprived areas; (b) population density quintile, with category 1 representing the least densely populated areas; (c) smoking (tobacco expenditure), with category 1 representing areas with the lowest tobacco expenditure; (d) NDVI quintile, with category 1 representing areas with the lowest NDVI values; (e) urban/rural classification; (f) ethnicity, percentage of residents recorded as White; and (g) regional boundaries. Population density, tobacco expenditure and NDVI were entered into the models as continuous variables and were grouped into quintiles here solely for visualisation.

Figure S2. Posterior median relative risks and 95% credible intervals estimated from models without adjustment for area-level covariates (left) and fully adjusted models (right).

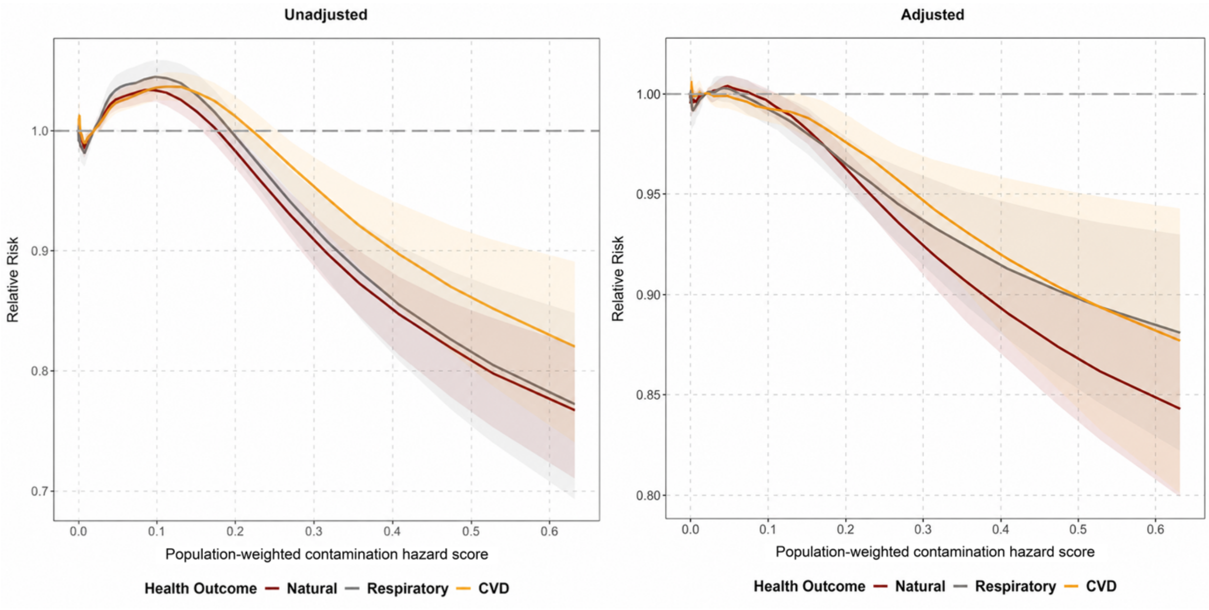


In models without adjustment for area-level covariates, while retaining the same spatial structure (left), the negative association at higher contamination hazard scores was still present, although the shape of the exposure-response curve differed from that of the fully adjusted models (right). The unadjusted model showed higher risks at low scores (around 0.08–0.18). These positive associations were attenuated after adjustment for income deprivation, population density, tobacco expenditure, green space, urbanicity, and ethnicity, while the negative associations at higher scores remained but were slightly weaker in magnitude.

Figure S3. Correlation between LSOA-level population-weighted contamination hazard score and continuous covariates.

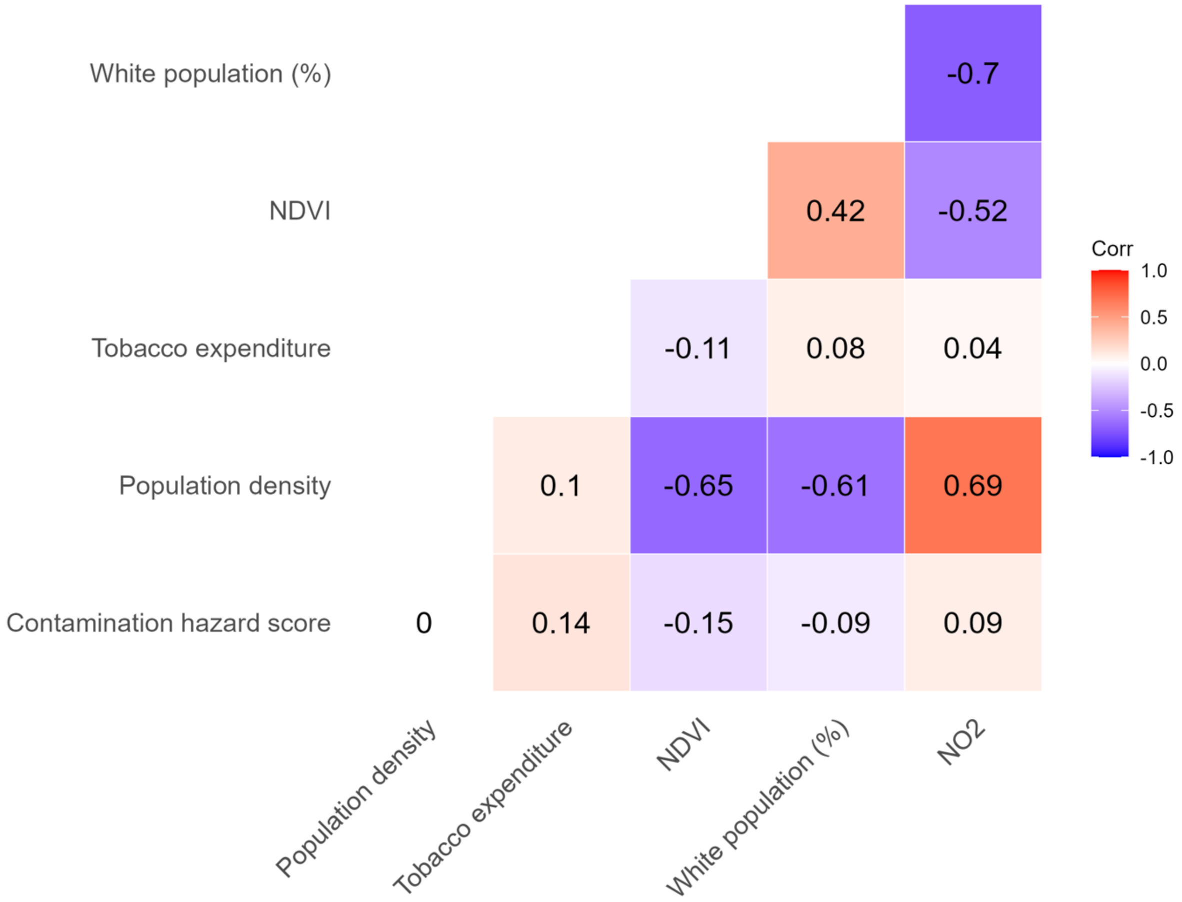

Figure S4. Distribution of population-weighted contamination hazard scores across LSOAs.

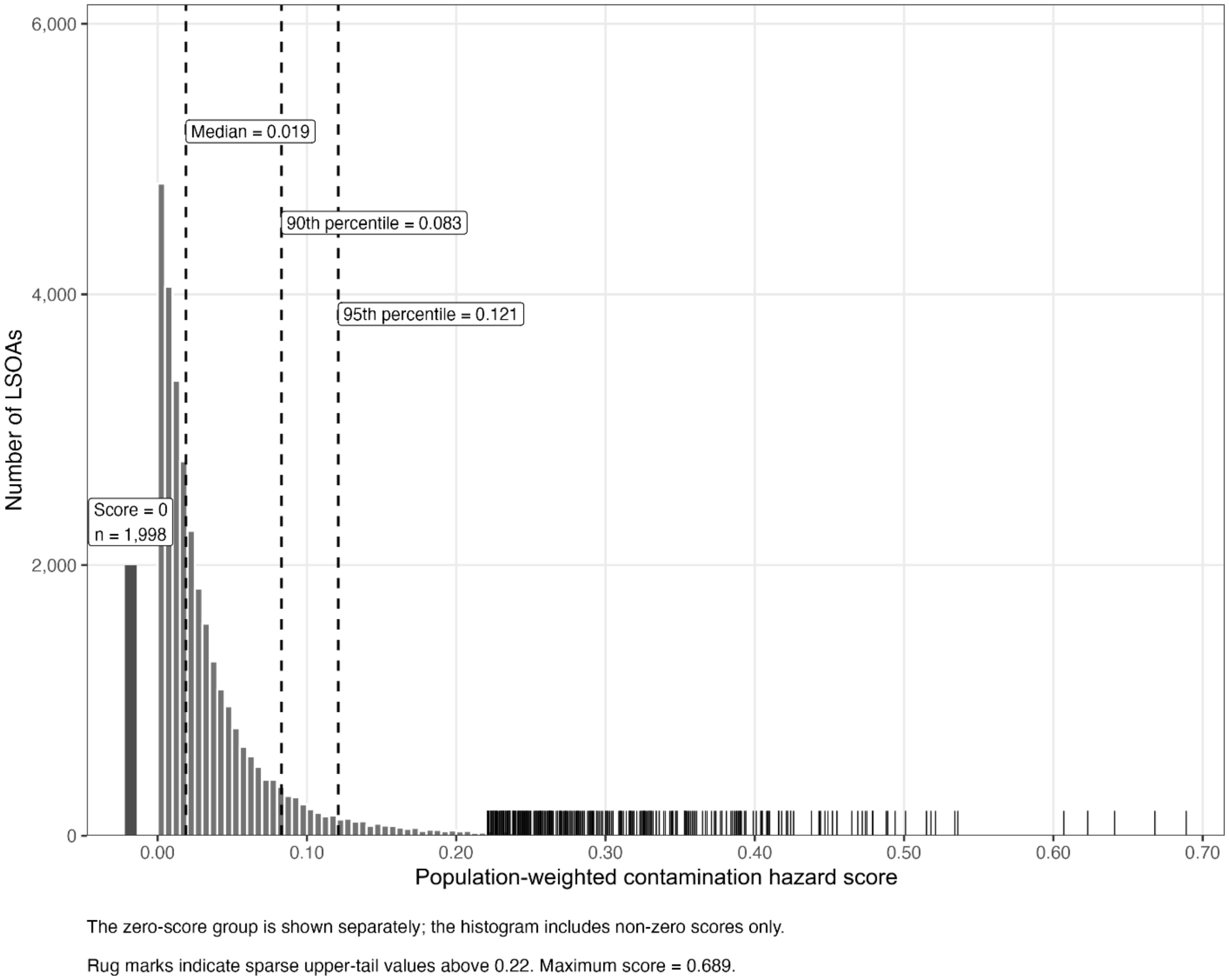


Distribution of the population-weighted contamination hazard scores across LSOAs. The zero-score group is shown separately and the histogram includes non-zero scores only. Vertical dashed lines show the 50$^{th}$ (median), 90$^{th}$, and 95th percentiles used in the sensitivity analyses. Rug marks show sparse upper-tail values above 0.22; the maximum score was 0.689.

Table S1. Distribution of continuous area-level covariates across LSOAs.

| Covariate | Median | IQR | Minimum | Maximum |
|---|---|---|---|---|
| **Tobacco expenditure (£/week/person)** | 4.72 | 3.88–5.84 | 1.49 | 12.77 |
| **Population density (persons/km²)** | 3,565 | 1,301–5,938 | 2 | 106,716 |
| **Non-White residents (%)** | 5.2 | 2.3–16.7 | 0 | 99.3 |
| **$NO_2$ concentration ($\mu g/m^3$)** | 20.25 | 16.53–24.36 | 5.16 | 64.42 |
| **NDVI** | 0.507 | 0.426–0.580 | −0.266 | 0.863 |

Notes: IQR, interquartile range ($25^{th}$–$75^{th}$ percentile interval). Statistics describe the unweighted distribution across 32,844 LSOAs. All variables have complete data. Quartiles were calculated using linear interpolation; population density is rounded to the nearest whole person/km².

Table S2. Sensitivity analysis using categories of the LSOA-level population-weighted contamination hazard score.

| **Category** | **Score range** | **n LSOAs** | **RR (95% CrI)** | | |
|---|---|---|---|---|---|
| | | | Natural-cause | Respiratory | CVD |
| **Four-category** | | | | | |
| **Zero** | 0 | 1,998 | 1.000 (reference) | 1.000 (reference) | 1.000 (reference) |
| **Low*** | 0.001–0.019 | 14,531 | 1.000 (0.994, 1.006) | 1.002 (0.992, 1.012) | 0.995 (0.987, 1.003) |
| **Moderate*** | >0.019–0.121 | 14,684 | 1.003 (0.997, 1.010) | 1.005 (0.995, 1.016) | 0.993 (0.985, 1.002) |
| **High*** | >0.121–0.689 | 1,631 | 0.975 (0.966, 0.984) | 0.982 (0.968, 0.997) | 0.980 (0.968, 0.992) |
| **Three-category** | | | | | |
| **Zero** | 0 | 1,998 | 1.000 (reference) | 1.000 (reference) | 1.000 (reference) |
| **>0 to $90^{th}$ percentile** | 0.001–0.083 | 27,582 | 1.001 (0.995, 1.008) | 1.004 (0.994, 1.014) | 0.995 (0.987, 1.003) |
| **>$90^{th}$ percentile** | >0.083–0.689 | 3,264 | 0.985 (0.977, 0.993) | 0.985 (0.972, 0.997) | 0.982 (0.972, 0.992) |

***Low category: contamination hazard score > 0 to the median; moderate category: >median to the $95^{th}$ percentile; high category: >$95^{th}$ perceentile.**

Note: RR, relative risk; CrI, credible interval; CVD, cardiovascular disease; LSOA, Lower-layer Super Output Area. Estimates are from fully adjusted Bayesian hierarchical spatial models accounting for area-level heterogeneity and spatial dependence. The four-category model used zero, the median, and the $95^{th}$ percentile as cut-points. The three-category model used zero and the $90^{th}$ percentile as cut-points.

Table S3. Sensitivity analysis: adjusted associations between the population-weighted contamination hazard scores and emergency hospital admissions, estimated using linear regression across 32,844 LSOAs.

| Model | Exposure | Natural | Respiratory | CVD |
|---|---|---|---|---|
| **Categorical** | Zero (reference) | | | |
| | >0 to 90$^{th}$ percentile | -0.023 (-0.030, -0.015) | -0.019 (-0.030, -0.007) | -0.019 (-0.029, -0.010) |
| | >90$^{th}$ percentile | -0.057 (-0.068, -0.045) | -0.040 (-0.057, -0.023) | -0.049 (-0.063, -0.035) |
| **Continuous** | Per IQR increase (log-transformed score) | -0.014 (-0.016, -0.012) | -0.009 (-0.013, -0.006) | -0.012 (-0.015, -0.009) |

To assess sensitivity to the modelling framework, we fitted ordinary least squares linear regression models of LSOA-level standardised hospital admission ratios (observed/expected admissions). Models included the same covariates as the main analysis (log population density, income quintile, tobacco expenditure, NDVI, rural-urban classification, % non-White ethnicity and region), with region entered as a fixed effect and no spatial random effects or exposure smoothing. Contamination hazard scores were modelled either using the three-category (zero [reference] and 90$^{th}$ percentile split) or as a continuous log-transformed term, with coefficients expressed per interquartile-range increase (1.781 log units).

Table S3 presents adjusted regression coefficients ($\beta$) with 95% confidence intervals from linear regression models. Compared with LSOAs with zero contamination hazard scores, coefficients for the moderate- and high-score categories were negative across all three outcomes, with larger differences in the high-score category. Continuous models also showed negative associations per interquartile-range increase in the log-transformed score. All 95% confidence intervals excluded zero.
